# Transition of antibody titers after the SARS-CoV-2 mRNA vaccine in Japanese healthcare workers

**DOI:** 10.1101/2021.12.28.21268435

**Authors:** Masahiro Kitabatake, Noriko Ouji-Sageshima, Shota Sonobe, Ryutaro Furukawa, Makiko Konda, Atsushi Hara, Hiroyasu Aoki, Yuki Suzuki, Natsuko Imakita, Akiyo Nakano, Yukio Fujita, Shigeyuki Shichino, Ryuichi Nakano, Satoshi Ueha, Kei Kasahara, Shigeo Muro, Hisakazu Yano, Kouji Matsushima, Toshihiro Ito

## Abstract

Since February 2021, health care workers in Japan have been preferentially vaccinated with a messenger RNA vaccine (BNT162b2/Pfizer) against severe acute respiratory syndrome coronavirus 2 (SARS-CoV-2). While many studies have confirmed that this vaccine is highly effective in reducing hospitalizations and deaths from coronavirus disease 2019 (COVID-19), antibody titers tend to decline at 3 months, leading to a risk of breakthrough infections. Thus, information is needed to support decision making regarding the third vaccination. In this study, we investigated transition of the anti-SARS-CoV-2 receptor-binding domain (RBD) IgG and neutralizing antibody titers of 41 vaccinated Japanese healthcare workers. Samples were collected seven times starting 1 week before vaccination until 6 months post-vaccination. Anti-SARS-CoV-2 RBD IgG levels peaked at 7 days after the booster, then declined over time and decreased to <10% at 6 months after the booster. Workers with low anti-SARS-CoV-2 RBD IgG levels also had low neutralizing antibody titers. These data support the active use of boosters for healthcare workers, especially for those with low anti-SARS-CoV-2 RBD IgG levels.

## Introduction

The efficacy and safety of the BNT162b2 mRNA coronavirus disease 2019 (COVID-19) vaccine (Pfizer-BioNTech, Mainz, Germany) has been widely demonstrated^1^. Real-world data on BNT162b2 have confirmed that this vaccine is highly effective in reducing laboratory-confirmed and community-acquired infections, viral loads in infected individuals, and COVID-19-related hospitalizations and deaths^2,3^. However, breakthrough infections have been reported in fully vaccinated persons^4^. Healthcare workers are potentially at a higher risk of infection owing to prolonged and repeated exposure to severe acute respiratory syndrome coronavirus 2 (SARS-CoV-2)-infected patients^5^. A recent study reported that among fully vaccinated health care workers, the occurrence of SARS-CoV-2 breakthrough infections was correlated with neutralizing antibody titers during the peri-infection period^3^. Another study found that patients’ humoral responses were substantially decreased 6 months after receiving the second dose of the BNT162b2 vaccine^6^. In this study, we evaluated the transition of antibody titers from the SARS-CoV-2 mRNA vaccine in 41 Japanese healthcare workers and the relationship between anti-SARS-CoV-2 receptor-binding domain (RBD) IgG levels and neutralizing antibody titers 6 months after the workers received the second BNT162b2 vaccine.

## Methods

Forty-one healthcare workers at Nara Medical University in Japan who had received the Pfizer-BNT162b2 vaccine between 10 March and 30 March 2021 were invited to participate as volunteers in the study. Participants provided peripheral blood samples for serologic assays before receiving the first vaccine, 1 week after receiving the first vaccine, then 1 week, 1 month, 3 months, and 6 months after receiving the second vaccine. All study participants provided written informed consent. Eligibility criteria were that participants were ≥20 years of age and had no history of suspected clinical SARS-CoV-2 infection. The Nara Medical University Ethics Committee approved the study (No. 3168).

All serum samples from vaccinated participants were tested for antibodies against the SARS-CoV-2 spike protein RBD with a COVID-19 human IgG enzyme-linked immunosorbent assay kit per the manufacturer’s instructions (RayBiotech Life, Inc. Peachtree Corners, GA, USA). The SARS-CoV-2 virus neutralization assay was performed as described previously^7^. SARS-CoV-2 (nCoV-19/JPN/TY/WK521/2020) was isolated and provided from the National Institute of Infectious Diseases, Japan. Serial 2-fold dilutions of heat-inactivated sera were incubated with equal volumes of 200 plaque-forming units of SARS-CoV-2 at 37°C for 1 h. VERO E6/TMPRSS2 cells (JCRB Cell Bank, Japan, JCRB1819) were infected with half the volume of the virus-serum mixture for 1 h, then covered with agarose overlay. After 48 h of incubation, cells were fixed and inactivated with 10% formalin, then the agarose overlay was removed, and the cells were stained with crystal violet. The visible plaques were counted, then 50% plaque-reduction neutralization tests were used to determine the titers as the maximum serum dilution that reduced the plaque number by >50%. All experiments using SARS-CoV-2 were performed in a biosafety level 3 facility at Nara Medical University, Japan.

## Results

We investigated the transition of anti-SARS-CoV-2 RBD IgG levels during the 6-month follow-up. The RBD IgG levels were slightly increased 7 days after participants received the first dose of the Pfizer-BNT162b2 vaccine (43.8 binding antibody units (BAU) /ml, 95% confidence interval [CI]: 15.3–125.4) compared with those before the vaccination (10.3 BAU/ml, 95% CI: 3.5–29.9). The highest titers were observed 7 days after the second dose of the Pfizer-BNT162b2 vaccine (4549 BAU/ml, 95% CI: 3604–5740), then the titers decreased by factors of 3.4 after 1 month (1341 BAU/ml, 95% CI: 1077–1671), 6.6 after 3 months (684.2 BAU/ml, 95% CI: 552.8–895.5), and 15.3 after 6 months (297.3 BAU/ml, 95% CI: 217.5–406.3).

Next, to investigate whether the anti-SARS-CoV-2 RBD IgG levels were correlated with the neutralizing antibody levels, we examined the neutralizing antibody values, expressed as 50% neutralizing titers, from the highest eight and lowest eight SARS-CoV-2 RBD IgG levels from participants 6 months after the second vaccination. The anti-SARS-CoV-2 RBD IgG levels were positively correlated with the neutralizing antibody levels, and low SARS-CoV-2 RBD IgG levels indicated very low or undetectable neutralizing antibody levels.

## Discussion

Studies are ongoing regarding SARS-CoV-2, long-term vaccine effectiveness, breakthrough infections, and effectiveness of the vaccine against mutant strains after the second dose of the BNT162b2/Pfizer vaccine. Our results indicated that BNT162b2 boosters generated large antibody responses in Japanese healthcare workers, but the anti-SARS-CoV-2 antibody titers decreased to approximately 1/15 at 6 months after the second vaccination. Low serum SARS-CoV-2 RBD IgG levels were strongly correlated with low neutralizing antibody levels against SARS-CoV-2. Healthcare workers are at a higher risk for SARS-CoV-2 infection, and the SARS-CoV-2 infection rate in healthcare workers reportedly varies between 3% and 17% according to the history, degree of exposure and presence of symptoms^8^. Therefore, in Japan, the BNT162b2 vaccination was recommended for healthcare workers in advance because of its high effectiveness in preventing confirmed infections, critical disease, and death. A recent study demonstrated substantially decreased humoral immunity against SARS-CoV-2 at 6 months after receiving the second dose of the BNT162b2 vaccine^6^. These data suggest that the booster should be highly recommended for healthcare workers, especially those with low serum SARS-CoV-2 RBD IgG levels 6 months after the second dose. Additionally, some reports indicate lower effectiveness against mutant SARS-CoV-2 strains, such as delta and omicron^9,10^.

This study had some limitations, including a small sample size, no use of mutant strains in the neutralization responses, and an absence of cellular immune responses. Additionally, other than for the humoral response, data are lacking regarding cell-mediated immune responses and to what extent this protection contributes to the long-term efficacy of the vaccine. Therefore, the effects of long-term transition and boosters on cell-mediated immunity should be investigated in detail.

**Figure 1.**
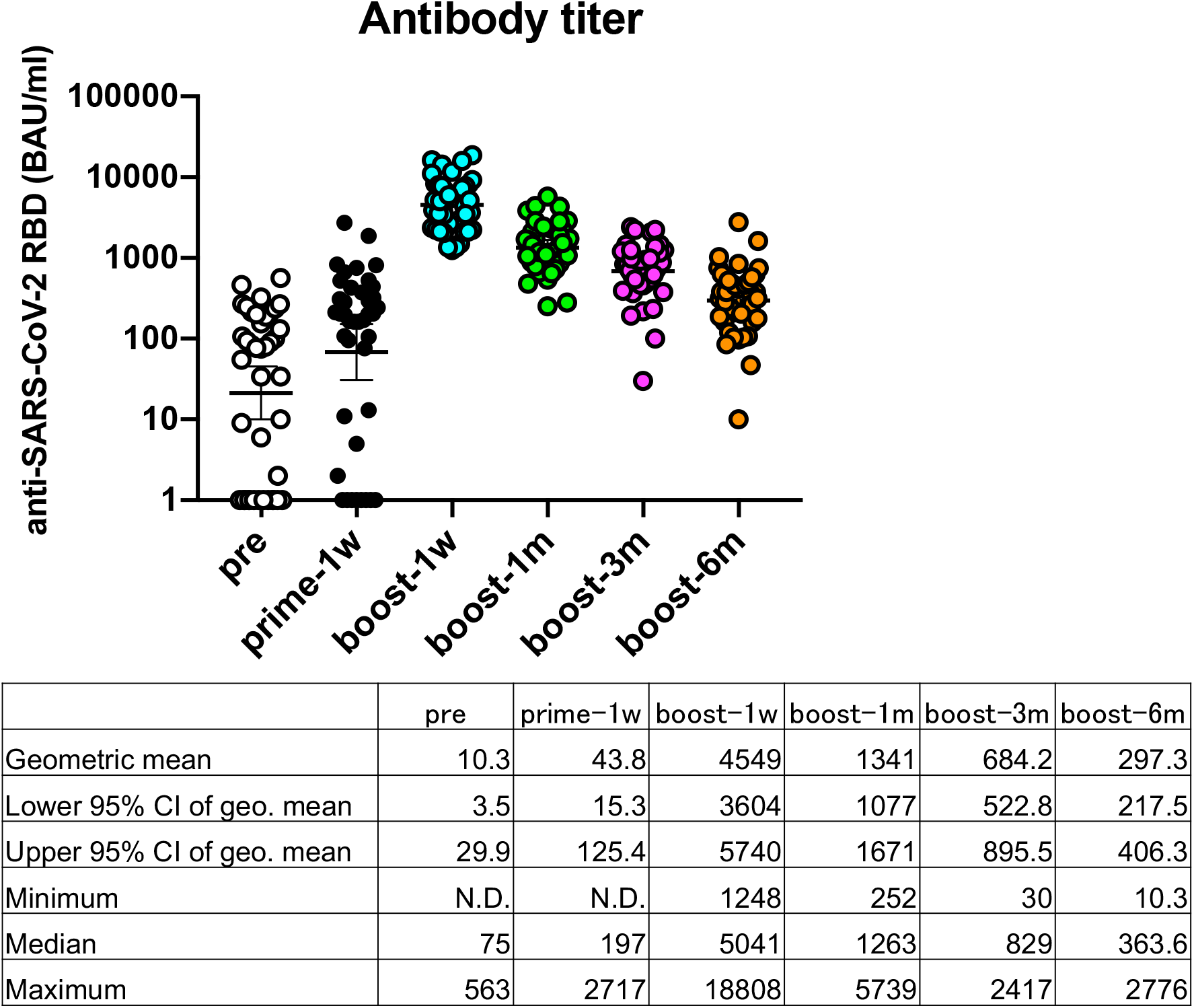
Transition of anti-SARS-CoV-2 responses after the COVID-19 mRNA BNT162b2/Pfizer vaccination. Responses to the COVID-19 mRNA vaccine were measured as anti-SARS-CoV-2 RBD IgG antibodies before the first vaccination (pre), 1 week after the first vaccination (prime-1w), and 1 week (boost-1w), 1 month (boost-1m), 3 months (boost-3m), and 6 months (boost-6m) after the second vaccination (n=41 participants). N.D.= Not detectable.

**Figure 2.**
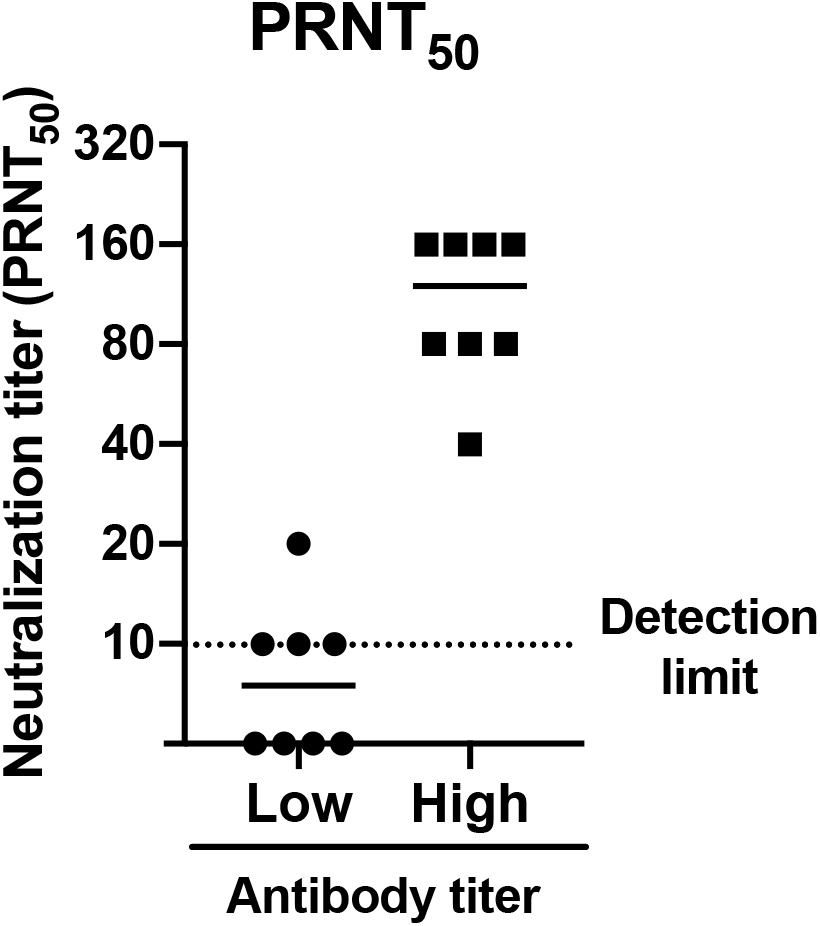
Distribution of the neutralizing antibody response 6 months after the COVID-19 mRNA booster. Neutralizing antibody levels from the eight highest and eight lowest anti-SARS-CoV-2 RBD antibody levels were measured via 50% plaque-reduction neutralization test (PRNT_50_) titers.

## Data Availability

All data produced in the present work are contained in the manuscript.

## Acknowledgments

We thank Ms. Hisayo Nishikawa (Department of Immunology, Nara Medical University), for their assistance. We also thank Traci Raley, MS, ELS, from Edanz (https://jp.edanz.com/ac) for editing a draft of this manuscript.

